# Investigating the clinico-anatomical dissociation in the behavioral variant of Alzheimer’s disease

**DOI:** 10.1101/19006676

**Authors:** Ellen H. Singleton, Yolande A. L. Pijnenburg, Carole H. Sudre, Colin Groot, Elena Kochova, Frederik Barkhof, Renaud La Joie, Howard J. Rosen, William W. Seeley, Bruce Miller, M. Jorge Cardoso, Janne Papma, Philip Scheltens, Gil D. Rabinovici, Rik Ossenkoppele

## Abstract

**Objective:** We previously found temporoparietal “Alzheimer-typical” atrophy in patients with the behavioral variant of Alzheimer’s disease (bvAD) with relative sparing of frontal regions. Here, we aimed to understand the pathophysiological mechanisms of bvAD based on alternative neuroimaging markers.

**Methods:** We retrospectively included 150 participants at the University of California San Francisco and University of Berkeley, including 29 bvAD, 28 “typical” amnestic-predominant AD (tAD), 28 behavioral variant of frontotemporal dementia (bvFTD), and 65 cognitively normal participants. Patients with bvAD were compared with other groups on glucose metabolism and metabolic connectivity on [^18^F]FDG-PET, and subcortical gray matter volumes and white matter hyperintensity volumes (WMHV) on MRI. A receiver-operating-characteristic-analysis was performed to determine the measures yielding the highest contrast between groups.

**Results:** bvAD and tAD showed predominant temporoparietal hypometabolism compared to controls, and did not differ in direct contrasts. However, overlaying statistical maps from contrasts between patients and controls revealed broader frontoinsular hypometabolism in bvAD compared to tAD, partially overlapping with bvFTD. Metabolic connectivity analyses indicated greater anterior default mode network (DMN) involvement in bvAD compared to tAD, mimicking bvFTD. Analyses of subcortical volume and WMHV showed no relevant group differences. The top-3 discriminative measures for bvAD *vs*. bvFTD were: metabolism in posterior (bvAD<bvFTD), anterior DMN (bvAD>bvFTD) and parietal cortex (bvAD<bvFTD; AUC: 0.80-0.91, p<0.01), while the top-3 discriminators for bvAD *vs*. tAD were amygdalar volume (bvAD>tAD), anterior DMN (bvAD<tAD) and salience network metabolism (bvAD<tAD; AUC: 0.66-0.75, p<0.05).

**Conclusion:** Subtle frontoinsular hypometabolism and anterior DMN involvement may underlie the prominent behavioral phenotype in bvAD.

## INTRODUCTION

Individuals with the behavioral variant of Alzheimer’s disease (bvAD) present early and prominent behavioral and personality changes, with AD as the primary etiology^1^. Case reports and small sample studies have suggested prominent frontal atrophy and pathology in bvAD patients^2-5^. The largest neuroimaging study to date in clinically defined bvAD patients revealed a prominent temporoparietal atrophy pattern with a relative lack of frontal atrophy^1^, questioning the neurobiological basis of the prominent behavioral deficits. The behavioral phenotype in these individuals might be explained better by complementary neuroimaging techniques. For example, functional measures such as glucose hypometabolic patterns or alterations in metabolic connectivity may be more sensitive than structural MRI^6^ and allow the assessment of large-scale networks rather than sole investigation of localized associations^7^. Furthermore, structural measures such as subcortical atrophy or white matter damage affecting frontosubcortical tracts have consistently been associated with neuropsychiatric symptoms^8, 9^. Exploring these neuroimaging features will enhance our neurobiological understanding of the prominently behavioral phenotype in bvAD. In addition, it may aid the often challenging differential diagnosis of bvAD versus ‘typical’ AD or the behavioral variant of frontotemporal dementia (bvFTD)^5, 10^, and potentially lead to more accurate diagnoses and patient management. We had two study objectives: (i) to increase our understanding of the relative lack of frontal atrophy in patients with the behavioral variant of AD through the assessment of multiple neuroimaging markers, and (ii) to identify the diagnostic accuracy of several neuroimaging, neuropsychological and neuropsychiatric measures in the differential diagnosis of bvAD *vs*. typical AD and bvFTD.

## METHODS

### Participants

We selected 29 bvAD patients from the University of California San Francisco (UCSF) AD Research Center (San Francisco, USA) who were included in our previous report on bvAD^1^, and had available structural MRI, FLAIR or FDG-PET neuroimaging measures. In the absence of consensus clinical criteria for the behavioral variant of AD, patients with bvAD were defined retrospectively by a group of behavioural neurologists (G.D.R., Y.A.L.P., P.S.) and neuropsychologists (R.O., J.H.K.) as patients with a diagnosis of bvFTD or “frontal variant AD” or a differential diagnosis of bvFTD *vs*. AD who had biomarker evidence for and/or autopsy confirmation of AD pathology^1^. Data availability differed per modality, as FDG-PET (n=19) and FLAIR data (n=15) were only available in a subset (see Table 1 for characteristics of the group with MRI data available and Supplementary Tables S1 and S2 for characteristics of the FDG-PET and FLAIR subsets). Patients with bvAD were matched with 28 tAD patients and 28 bvFTD patients, also described in the original study^1^. tAD patients fulfilled criteria for probable AD with at least an intermediate-likelihood of AD pathophysiology according to the National Institute on Ageing-Alzheimer’s Association criteria^11^ or mild cognitive impairment due to AD^12^ based on positive amyloid biomarkers and/or autopsy. bvFTD patients met the clinical criteria proposed by Neary and colleagues^13^ or Rascovsky and colleagues^14^, and had negative amyloid biomarkers and/or autopsy confirmation. Patients with significant cerebrovascular disease were excluded from the UCSF Aging and Dementia Research Cohort. Finally, we selected two cognitively normal control groups. The first group underwent MRI on the same scanners as the patient groups at UCSF, but had no FDG-PET data available (CN_1_, n=34). The second group underwent FDG-PET on the same scanners as the patient groups at the University of California Berkeley (CN_2_, n=31), but had MRI on a different scanner than the patient groups. Both CN groups had cognitive test scores within the normal range and absence of neurological or psychiatric illness^15^.

**Table 1.** Demographic, neuropsychological and neuropsychiatric characteristics across groups. Differences between groups were assessed using ANOVA tests, Chi-square tests and Kruskall-Wallis tests with post hoc Mann-Whitney U-tests, where appropriate. All p-values were corrected for multiple comparisons using Bonferroni correction. Data presented above are based on the groups for whom T1 MRI scans were available. See Supplementary Tables S1 and S2 for equivalent information in groups for which FDG and FLAIR scans were available.

|  | bvAD | tAD | bvFTD | CN<sub>1</sub> | p-value |
| --- | --- | --- | --- | --- | --- |
| n | 29 | 28 | 28 | 34 |  |
| Age, y | 64.4 (9.4) | 63.0 (9.3) | 64.6 (4.4) | 64.9 (9.9) | 0.84 |
| Sex, no. male (%) | 17 (59) | 16 (55) | 21 (70) | 22 (65) | 0.82 |
| Education<sup>a</sup>, y mean (SD) | 15.7 (2.6) | 15.8 (2.8) | 15.1 (3.4) | 17.9 (2.0) | 0.001 |
| MMSE<sup>b†</sup>, mean (SD) | 22.0 (5.9) | 22.1 (5.7) | 21.3 (6.7) | 29.5 (0.7) | 0.001 |
| APOEε4 positivity<sup>c</sup>,<br>no. of patients (%) | 11/18 (61) | 10/14 (67) | 3/27 (11) | 6/34 (18) | <0.001 |
| MRI scanner field strength |  |  |  |  | 0.16 |
| 1.5T | 17 (59) | 22 (79) | 14 (50) | 22 (65) |  |
| 3T | 12 (41) | 6 (21) | 14 (50) | 12 (35) |  |
| Memory domain z-score<sup>d°</sup>,<br>mean (SD) | -3.5 (1.5) | -3.9 (1.3) | -2.8 (1.7) | 0.3 (0.9) | <0.001 |
| Executive domain z-score<sup>e°</sup>,<br>mean (SD) | -1.9 (1.0) | -1.9 (1.0) | -2.1 (1.0) | 0.0 (0.6) | <0.001 |
| NPI score<sup>f◇</sup>, mean (SD) | 30.2 (20.6) | 12.8 (15.4) | 34.7 (17.2) | - | 0.001 |

<sup>†</sup> MMSE data was available for n=26 for bvAD, n=19 for tAD, n=27 for bvFTD and n=34 for CN<sub>1</sub>
<sup>°</sup> Cognition data was available for n=22 for bvAD, n=28 for tAD, n=24 for bvFTD, n=30 for CN<sub>1</sub> and tested with MANOVA with Bonferroni correction.
<sup>◇</sup> NPI data was available for n=13 for bvAD, n=18 for tAD, n=20 for bvFTD and n=0 for CN<sub>1</sub>
<sup>a</sup> Controls > patients, p<0.01
<sup>b</sup> Controls > patients, $p < 0.001$
<sup>c</sup> bvAD & tAD > controls, $p < 0.01$ , bvAD & tAD > bvFTD, $p < 0.001$
<sup>d</sup> Controls > patients, $p < 0.001$ , tAD < bvFTD, $p < 0.05$
<sup>e</sup> Controls > patients, $p < 0.001$
<sup>f</sup> bvAD & bvFTD > tAD, $p < 0.01$

### Standard protocol approvals, registrations and patient consents

Informed consent was obtained from all subjects or their assigned surrogate decision-makers, and the study was approved by the University of California Berkeley, the UCSF and the Lawrence Berkeley National Laboratory institutional review boards for human research.

### Data availability statement

Anonymized data used in the present study may be available upon request to the corresponding author.

### Investigating the pathophysiological mechanisms in bvAD

#### Glucose hypometabolism

FDG-PET images were obtained at Lawrence Berkeley National Laboratory (LBNL) using a Siemens ECAT EXACT HR PET (n_bvAD_=15) or Biograph PET/CT (n_bvAD_=4) scanner. Acquisition parameters have been specified elsewhere^16^. Starting 30 min post-injection of 5-10 mCi of [^18^F] Fluorodeoxyglucose (FDG), 6 x 5 minutes frames of emission data were collected. All PET data were reconstructed using an ordered subset expectation maximization algorithm with weighted attenuation. Images were smoothed with a 4 × 4 × 4-mm Gaussian kernel with scatter correction. FDG-PET frames of 30-60 minutes post-injection aligned to the first frame and averaged. Next, each frame was realigned to the resultant mean image. These native space images were summed and standardized uptake value ratios (SUVr) were calculated by normalizing the summed FDG images to the mean activity in the pons, as glucose metabolism in this region has been shown to be preserved in AD^17^. A mutual information affine registration was used to coregister these normalized FDG-PET images to the corresponding MR image in native space. For the cognitively normal group with FDG-PET scans available (CN_1_), MRI scans were obtained on a 1.5T Magnetom Avanto System scanner (Siemens Inc., Iselin, NJ) at the University of Berkeley, with a 12-channel head coil run in triple mode. These images were used for PET processing only. Subsequently, the MRI images were registered to Montreal Neurological Institute (MNI) space and the FDG-PET images were then also transformed to MNI space using the individual deformation fields obtained from the coregistered MRI normalization. The normalized FDG-PET images were then smoothed using a 12-mm Gaussian kernel^18^. All images were visually inspected and deemed suitable for further analyses. Then, voxel-wise comparisons of FDG-SUVr images were performed in SPM12 (Welcome Trust Center for Neuroimaging, University College London, www.fil.ion.ucl.ac.uk/spm), using an analysis of covariance model that included age and sex as covariates. Pairwise contrasts were performed among the four groups (i.e. bvAD, tAD, bvFTD and CN_1_), which yields *T*-maps signifying the difference in SUVr for each voxel. For comparisons between patients and controls, we thresholded *T*-maps at p<0.05, family-wise error corrected at the voxel level, and an extent threshold of k=50 voxels. For contrasts between patient groups we applied an uncorrected threshold of p<0.001 and extent threshold of k=50 voxels due to smaller expected differences between groups. This yields binary maps of significant voxels for each comparison and we overlaid these maps for patients *vs*. control contrasts on an MNI brain template to visualize regional differences and overlap between groups.

#### Metabolic connectivity

Resting-state metabolic connectivity was examined in all groups using a voxel-wise interregional correlation analysis (IRCA) of FDG-PET data^19^. This method involved several steps^20^; i) selection of relevant networks, ii) definition of seed regions-of-interest (ROI) within key regions in these functional networks as described in previous literature, iii) generation of covariance maps by correlating the mean FDG-SUVr in the seed ROI with the mean FDG-SUVr in all voxels across the brain, and iv) comparing these covariance maps to functional network templates and calculating goodness-of-fit (GOF) scores for each network. For step (i), we selected networks from the literature that are thought to play a pivotal role in bvFTD and tAD^21^, including the default mode network (DMN)^22^, salience network (SN)^23^ and executive control network (ECN)^24^. To study the specific contribution of posterior *vs*. anterior DMN, the DMN was fractioned into anterior and posterior subsystems in accordance with previous studies^25-27^. For step (ii), the left posterior cingulate cortex (PCC, MNI coordinates: x=-8, y=- 56, z=26^25^) was selected as the seed region for the posterior DMN, the left anterior medial prefrontal cortex (amPFC, x=6, y=52, z=-2^25^) for the anterior DMN, the right frontoinsula (riFI, x=36, y=18, z=4^28^) for the SN, and the right dorsolateral prefrontal cortex (riDLPFC, x=44, y=36, z=20^24^) for the ECN. Spheres of 4mm were drawn around the abovementioned coordinates and, for each subject, mean FDG SUVr values were extracted from each of these spheres using Marsbar while using a gray matter mask to exclude PET counts from white matter and cerebrospinal fluid. For step iii), multiple linear regressions were performed in SPM12 to assess correlations between FDG uptake in each seed ROI and FDG uptake across the brain, resulting in interregional covariance maps. As the PET covariance analyses explored correlations between the seed region and each voxel *across subjects*, one interregional correlation map was obtained per group. The interpretation of these maps is based on the notion that regions covarying in levels of metabolism are associated to each other. Separate models were used for each group, resulting in five interregional covariance maps per group. These analyses were adjusted for age and sex. For step iv) the goodness-of-fit of the interregional covariance maps with standard functional network templates, published by the Stanford Functional Imaging in Neuropsychiatric Disorders Lab^29^ was assessed. As previously described^29^, these standard functional network templates were created by applying FSL’s MELODIC independent component analysis software to resting state fMRI data of 15 healthy control subjects. The network templates were downloaded as binary ROIs from http://findlab.stanford.edu/functional_ROIs.html. Goodness-of-fit was assessed by calculating the difference between the mean *T*-score of all voxels of the interregional covariance map (transformed SPM *T*-maps) inside the functional network template (*T*_inside_) and the mean *T*-value of all voxels outside the functional network template (*T*_outside_), i.e. goodness-of-fit = *T*_inside_ – *T*_outside_^30^. A high goodness-of-fit score indicated a high correspondence of the pattern of correlated regions based on similar FDG uptake with certain network architecture. Due to the group-level nature of these analyses, no statistics were performed on the GOF scores. In order to test the robustness of the goodness-of-fit between the covariance maps and the functional network templates, these analyses were repeated with an independent set of network templates from functional MRI data from 1000 healthy subjects as part of the Neurosynth project (http://neurosynth.org^31^). The templates were obtained by entering the MNI coordinates and downloading the generated functional networks. The templates were thresholded at a default threshold of *r*=0.2 using FSL to create binary masks.

#### Subcortical atrophy

We compared bvAD patients with tAD, bvFTD and CN groups on gray matter volumes of several subcortical structures, including the amygdala, nucleus accumbens, caudate nucleus putamen, globus pallidus, hippocampus and thalamus. Volumes were extracted from T1-weighted MR scans, obtained at the University of California San Francisco, either on a 1.5T (Magnetom Avanto System/Magnetom VISION system, Siemens, Erlingen, Germany, n_bvAD_=17) or 3T (Tim Trio, Siemens, Erlingen, Germany, n_bvAD_=12) all with a standard 12-channel head matrix coil. Acquisition parameters have been published previously^20^. Subcortical parcellations were performed using FSL FIRST^32^. First, the T1 images were transformed to MNI space using affine registration, and a subcortical mask was applied to the images. Next, subcortical structures were segmented bilaterally based on shape models and voxel intensities. All images were inspected visually after registration and segmentation. For each subcortical structure, left and right absolute volumes were generated, calculated in cm^3^, and grouped together in the analysis, as there were no volume differences based on laterality. Statistical differences in volumes between groups were assessed using a general linear model, including all subcortical structures, with age, sex, scanner field strength and total intracranial volume, which was obtain by summing the gray matter, white matter and CSF volumes after segmentation in SPM12^33^, as covariates. Significant group differences were indicated by p<0.05, Bonferroni corrected.

#### White matter hyperintensity volumes

Next, we compared bvAD patients with tAD, bvFTD and CN groups on white matter hyperintensity volumes (WMHV), using a Bayesian Model Selection (BaMoS) algorithm on FLAIR MR images^34, 35^. Briefly, this method is a hierarchical, fully unsupervised model selection framework based on a Gaussian mixture model for neuroimaging data which enables the distinction between different types of abnormal image patterns without a priori knowledge, accounting for observation outliers and incorporating anatomical priors. Lesion volumes were calculated for four equidistant concentrical regions of white matter between the ventricles and cortices per lobe bilaterally^36^. All FLAIR images were visually inspected prior to their inclusion in the algorithm and those with significant motion or reconstruction artifacts were excluded. The WMHV segmentation was checked for quality and images with evident over- or underestimation were re-analyzed with an adjusted algorithm until satisfactory segmentation was obtained. Regional WMHV were normalized to the population of cognitively normal subjects, and statistical differences in WMHV between groups were assessed using a generalized linear model with gamma probability distribution and log link, adjusting for age, sex, scanner field strength and total intracranial volume. Significant group differences were indicated by p<0.05 and no correction for multiple comparisons was used due to the large correlation between dependent variables.

### Investigating the diagnostic accuracy of multiple neuroimaging markers for differentiating bvAD from tAD and bvFTD

To aid clinical differential diagnosis, receiver operating characteristic (ROC) analyses were performed to examine the area-under-the-curve (AUC) for discriminating bvAD from tAD and bvFTD. As input for the AUC analysis we used various neuroimaging measures investigated in aim 1: measures of glucose metabolism, metabolic connectivity, subcortical atrophy and white matter hyperintensities. For glucose metabolism, we extracted SUVr values from two AD-signature ROIs (i.e. temporoparietal cortex^37^ and a total parietal ROI based on the Automated Anatomical Labeling (AAL) atlas regions^38^) and one FTD-signature ROI (comprising the anterior cingulate, frontoinsular, striatal and frontopolar AAL atlas regions^39, 40^). In order to create individual measures of metabolic connectivity, we extracted mean SUVr values within the functional network templates as provided by Shirer et al. (2012) and divided them by the SUVr values outside the network templates (SUVr_within network_/SUVr_outside network_), thereby creating individual goodness-of-fit ratios based on SUVr values. For the subcortical structures, only the amygdala was added to the AUC analysis based on assessment of differences in subcortical volumes between diagnostic groups. For white matter hyperintensity volumes, total WMHV were included, without correction for total intracranial volume. Since we were interested in how the aim 1 neuroimaging measures related to structural MRI measures, we additionally used relevant structural MRI as input for the ROC analyses. We extracted gray matter volumes from the same AD-signature and FTD-signature ROIs as used for glucose metabolism analyses. Pairwise ROC analyses between all groups were performed separately for all measures, as the sizes of the groups per modality varied. We present the top-5 best discriminatory variables in the main text, and provide an overview of all results in Supplementary Table S7. In a sensitivity analysis, the ROC analyses were repeated including only patients that had both FDG-PET and MRI available.

## RESULTS

### Demographic characteristics

The patient groups did not differ in age, sex and MMSE (Table 1). The level of education was higher in the cognitively normal group compared to each of the three patient groups (p<0.01), while there were no differences between the patient groups. The proportion of APOEε4 positive patients was higher in the bvAD and tAD groups compared to cognitively normal controls (p<0.01) and bvFTD patients (p<0.001). Cognitive and NPI scores are presented in Table 1. There were no substantial differences in demographic characteristics between the different subsets of patients that had FDG-PET, MRI or FLAIR available (see Supplementary Tables S1 and S2).

### Investigating the pathophysiological mechanisms in bvAD

#### Glucose hypometabolism

Compared to cognitively normal controls, marked hypometabolism was found in the posterior cingulate, precuneus and lateral temporoparietal regions in both bvAD and tAD, while bvFTD displayed hypometabolism mainly in frontal regions and the temporal poles (Figure 1A&B). In direct patient group contrasts, bvAD showed no differences in glucose metabolism with tAD and less frontal hypometabolism than bvFTD (p<0.001, uncorrected, see Figure S1 for spatial patterns of patient *vs*. patient contrasts). Visual assessment of the overlay of *T*-maps resulting from voxel-wise comparisons between patients and cognitively normal controls suggested broader frontoinsular involvement in bvAD than in tAD, comprising the right lateral frontal lobe and bilateral insulae (Figure 1C).

**Figure 1.**
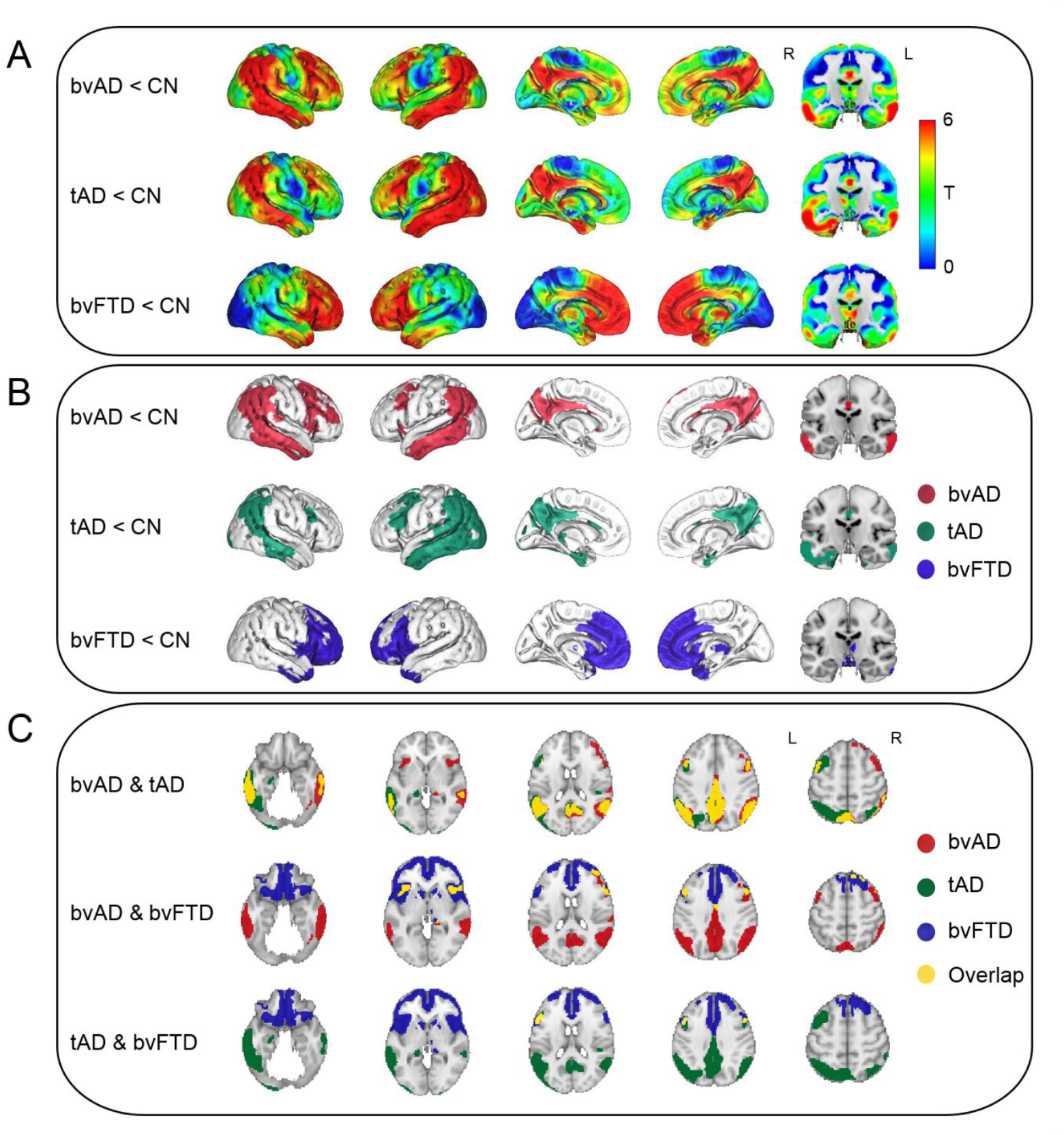
Patterns of hypometabolism of patients versus cognitively normal controls. *Panel A*) Surface rendering of *T*-maps showing hypometabolic regions in patient groups compared to cognitively healthy controls. Contrasts were adjusted for age and sex. *Panel B*) Surface rendering of significant voxels from contrasts between patients and controls, displayed at p<0.05, family-wise error corrected, extent threshold k=50. *Panel C*) Overlay of the *T*-maps from the voxel-wise comparison of FDG-PET SUVr between patients and controls. Overlays are displayed at p<0.05, family-wise error corrected, extent threshold k=50. Cerebellum was removed for visualization purposes.

#### Metabolic connectivity

bvAD patients showed a higher GOF score in the anterior DMN than tAD patients (*GOF*=4.13 versus 2.92, respectively), which was identical to the bvFTD GOF score (4.13). The GOF score for bvAD (3.85) in the posterior DMN was intermediate between bvFTD (2.04) and tAD (4.14), but closer to tAD. In the salience network, bvFTD had a higher GOF score (2.90) than both tAD (0.62) and bvAD (1.05). In the executive control network, bvAD patients (2.20) showed a lower GOF score than tAD (3.11) and bvFTD (2.78) patients (Figure 2 & Supplementary Table S4). Sensitivity analyses using different functional network templates showed a similar pattern of GOF scores (see Supplementary Table S4 & Figure S2).

**Figure 2.**
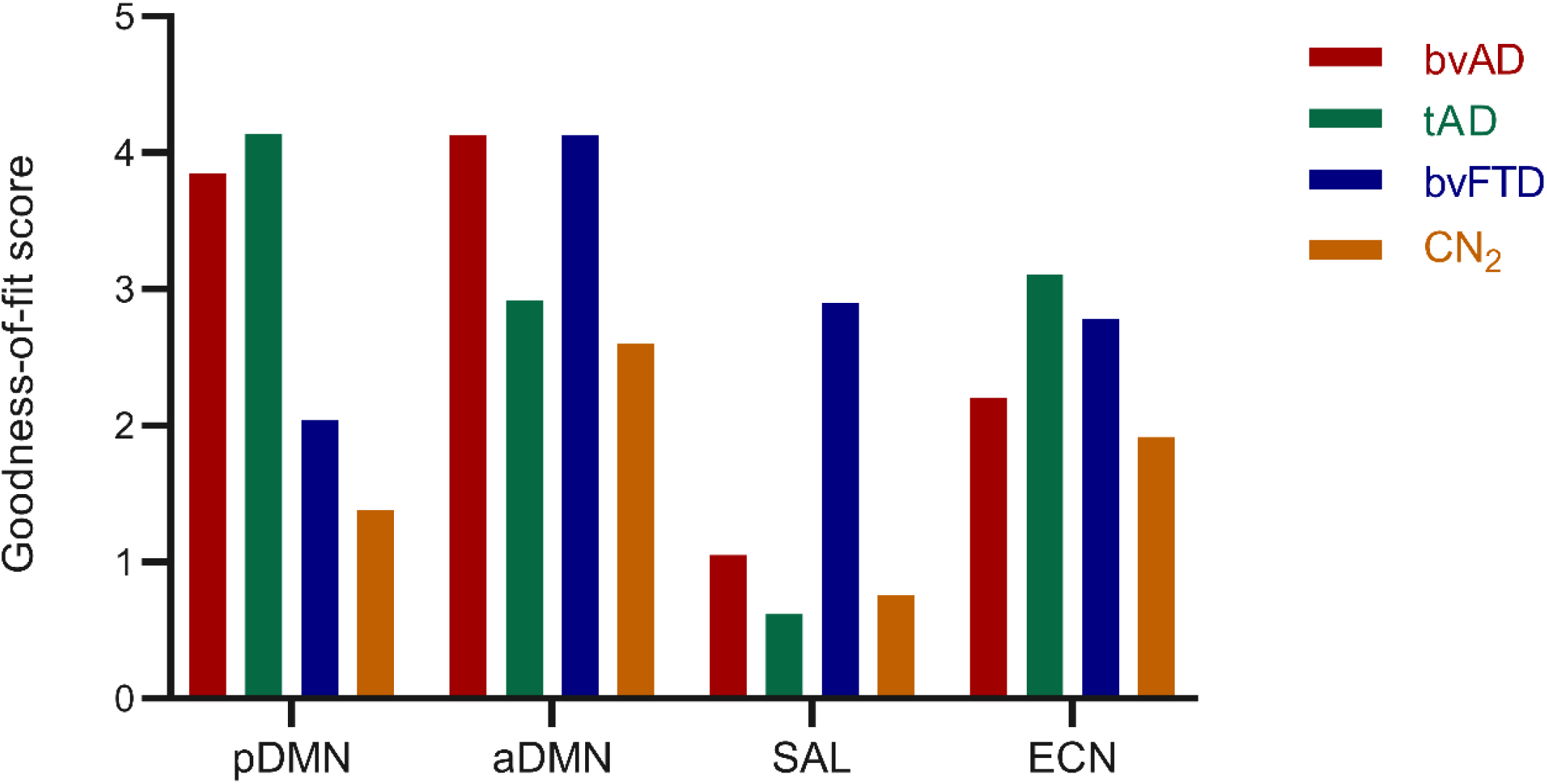
Goodness-of-fit (GOF) scores indicatng the resemblance of the FDG interregional covariance maps with intrinsic functional connectivity network templates. GOF scores represent the subtraction of the mean *T*-score outside of the network template from the mean *T*-score within the network template. *pDMN* = posterior default mode network, *aDMN* = anterior default mode network, *SAL* = salience network, *ECN* = executive control network.

#### Subcortical atrophy

Compared to cognitively normal controls, bvAD showed lower gray matter volumes in the hippocampus, putamen, caudate nucleus and thalamus, and no significant differences in the amygdala, nucleus accumbens and globus pallidus, while tAD patients showed lower volumes in the hippocampus, amygdala, nucleus accumbens and thalamus and bvFTD patients showed lower volumes in all subcortical structures compared to cognitively normal controls. bvAD showed larger amygdala gray matter volume than tAD (p<0.05) and no differences with tAD in all other examined structures. In comparison with bvFTD, bvAD and tAD patients showed larger globus pallidus gray matter volumes (p<0.05), tAD patients showed larger nucleus accumbens gray matter volumes (p<0.05), and no differences with bvAD were found in other structures (Figure 3 and Supplementary Table S5).

**Figure 3.**
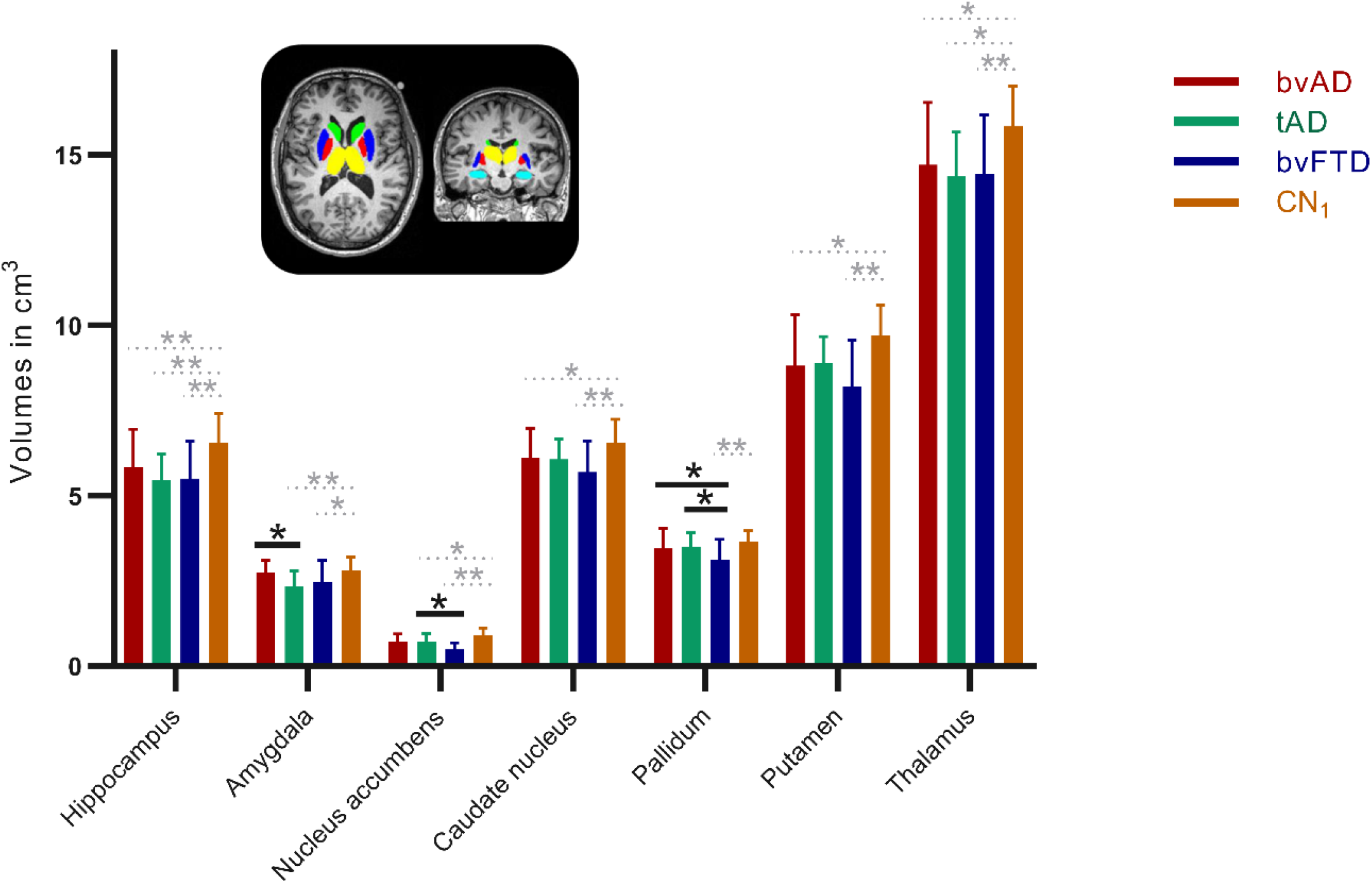
Gray matter volumes of subcortical structures across diagnostic groups. Volumes are displayed in cm^3^. Error bars indicate standard deviations. *p<0.05, **p<0.001, Bonferroni corrected (black indicating patient contrasts, while gray represents patient vs control contrasts). Green structures indicate the caudate nucleus, dark blue structures indicate the putamen, red structures indicate the globus pallidus, yellow indicate the thalamus, and light blue structures indicate the amygdala.

#### White matter hyperintensities

No differences were found between the patient groups in total WMHV, nor were regional differences found in WMHV between the patient groups (all p>0.05, Supplementary Table S6). Subregional analysis revealed lower frontal juxtacortical WMHV in bvAD than bvFTD, as well as lower left temporal juxtacortical WMHV, and higher right temporal juxtacortical WMHV in bvAD than bvFTD (p<0.05, Figure 4). In comparison to tAD, bvAD patients showed lower juxtacortical left temporal and subcortical WMHV and higher right temporal juxtacortical WMHV (p<0.05, Figure 4).

**Figure 4.**
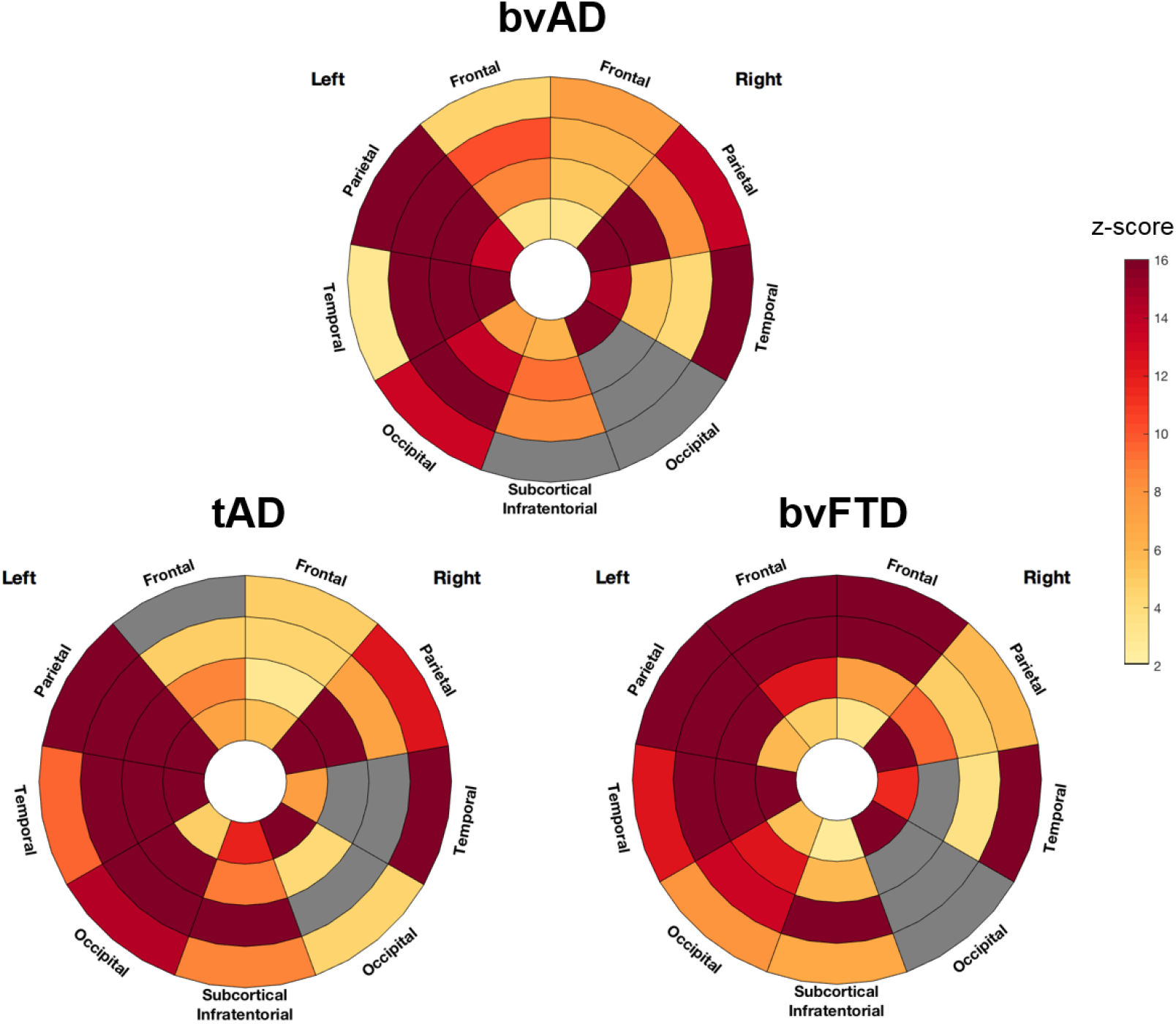
Regional distribution of white matter hyperintensity volumes in patient groups. In this plot, the angular sections correspond to different lobes while the concentric rings represent equidistant layers of white matter. Radius increases with the distance to the ventricles (center layer: periventricular – outer layer: juxtacortical). Grayed-out regions indicate regions where the difference when compared to the control group did not reach significance. Colored regions from light yellow to red indicate the multiplicative factor when compared to control group after correction for sex, scanner type and total intracranial volume.

### Investigating the diagnostic accuracy of multiple neuroimaging for differentiating bvAD from tAD and bvFTD

The top-5 discriminative variables for bvAD *vs*. bvFTD were posterior DMN (bvAD < bvFTD; AUC=0.91, 95%CI= 0.80-1.00) and anterior DMN (bvAD > bvFTD; AUC=0.83, 95%CI=0.70-0.97) metabolism, parietal metabolism (bvAD < bvFTD; AUC=0.80, 95%CI=0.65-0.94), salience network metabolism (bvAD > bvFTD; AUC=0.76, 95%CI=0.60-0.92), and temporoparietal metabolism (bvAD < bvFTD; AUC=0.75, 95%CI=0.58-0.91; Figure 5 & Table S8). bvAD was discriminated best from tAD by amygdala gray matter volume (bvAD > tAD; AUC=0.75, 95%CI=0.62-0.88), anterior DMN metabolism (bvAD < tAD; AUC=0.71, 95%CI=0.54-0.89), salience network metabolism (bvAD < tAD; AUC=0.66, 95%CI=0.49-0.84), FTD-signature region metabolism (bvAD < tAD; AUC=0.65, 95%CI=0.46-0.83) and FTD signature region gray matter volume (bvAD < tAD; AUC=0.61, 95%CI=0.46-0.76). The top-5 discriminative variables for bvAD *vs*. CN were temporoparietal hypometabolism (AUC=0.93, 95%CI=0.86-1.00), parietal hypometabolism (AUC=0.91, 95%CI=0.83-1.00), temporoparietal atrophy (AUC=0.91, 95%CI=0.83-0.99), hypometabolism in the posterior DMN (AUC=0.90, 95%CI=0.79-1.00) and parietal atrophy (AUC=0.89, 95%CI=0.79-0.99). A summary of all measures is included in Supplementary Table S7. Repeating the analyses with individuals having both MRI and FDG-PET data available showed similar results (see Supplementary Table S9 and Supplementary Figure S3).

**Figure 5.**
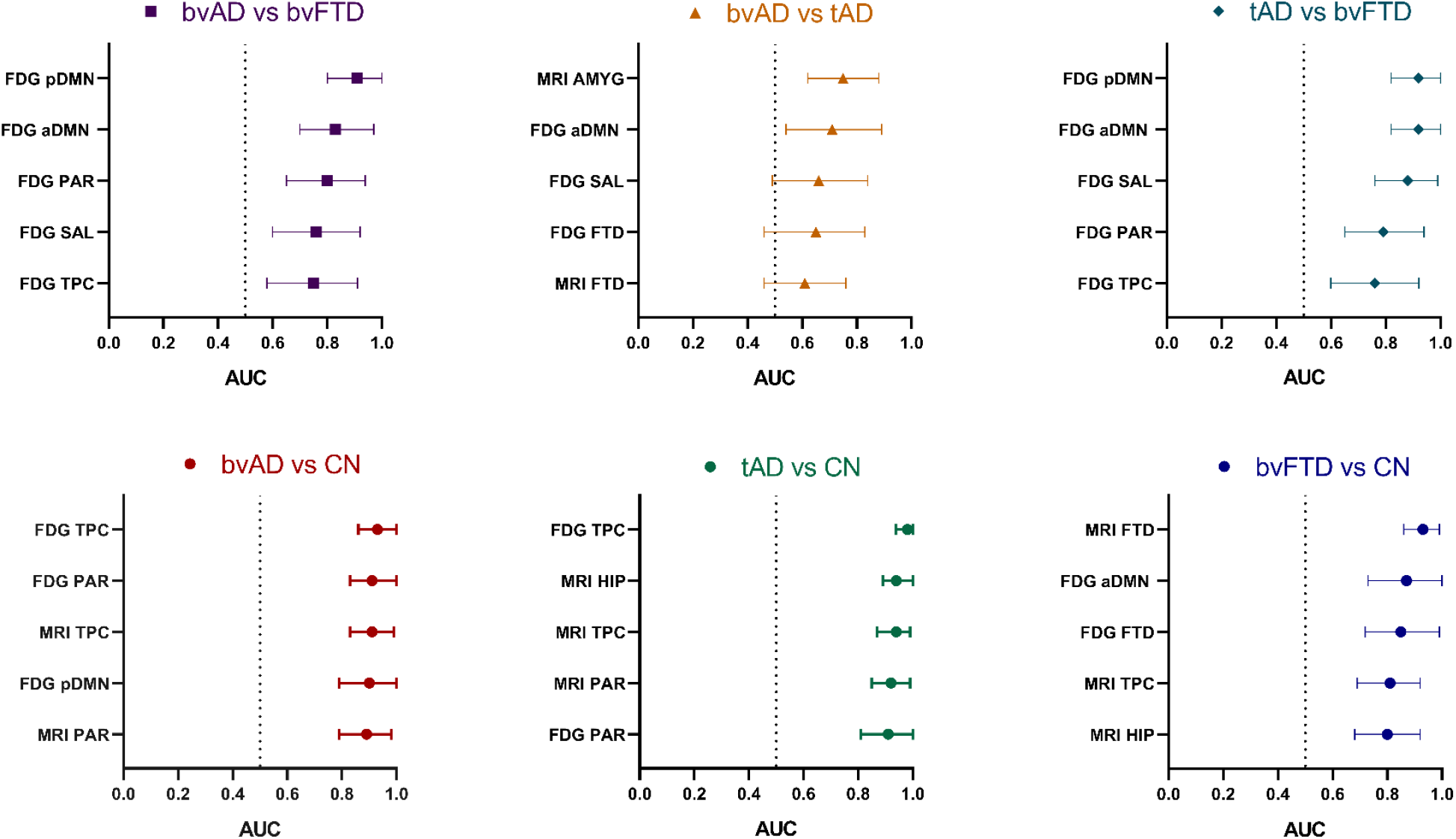
Top-5 discriminators for each contrast. The area-under-the-curve (AUC) and its 95% confidence interval are presented. *MRI FTD* = FTD signature region gray matter volume on MRI, consisting of the anterior cingulate, frontoinsula, striatum and frontopolar regions^39, 40^; *MRI AMYG* = bilateral amygadala gray matter volume on MRI; *MRI HIP* = bilateral hippocampus gray matter volume; *MRI PAR =* parietal gray matter volume; *MRI TPC* = temporoparietal gray matter volume; *FDG TPC* = temporoparietal cortex metabolism on FDG-PET; *FDG PAR* = parietal cortex metabolism on FDG-PET; *FDG pDMN* = glucose metabolism within the posterior default mode network, divided by the glucose metabolism without the posterior default mode network on FDG-PET; *FDG aDMN* = glucose metabolism within the anterior default mode network, divided by the glucose metabolism without the anterior default mode network on FDG-PET; *FDG SAL* = glucose metabolism within the salience network, divided by the glucose metabolism without the salience network on FDG-PET.

## DISCUSSION

The aims of the current study were (i) to explore the clinico-anatomical dissociation observed in bvAD (i.e. relative lack of frontal atrophy with prominent behavioral changes^1^) through assessment of multiple imaging markers and (ii) to examine the diagnostic accuracy of several neuroimaging, neuropsychological and clinical measures for differentiating bvAD from tAD and bvFTD. We hypothesized that bvAD patients would exhibit more anterior hypometabolism, more pronounced disintegration of metabolic connectivity networks involved in behavioral processes, greater subcortical atrophy and a greater white matter hyperintensity burden in regions impacting frontosubcortical tracts compared to tAD, and would partly resemble the neuroimaging characteristics of bvFTD. Our results suggest that the behavioral symptoms presented by bvAD patients are associated with subtle frontoinsular hypometabolism and increased anterior default mode involvement. In addition, our results suggest that subcortical atrophy and white matter hyperintensities do not contribute substantially to the clinical phenotype of bvAD. The ROC analysis reinforces the idea that the differentiation of bvAD from tAD and bvFTD lies in functional imaging rather than structural markers.

The pattern of hypometabolism in bvAD points towards subtle loss of neural activity in frontoinsular regions in addition to posterior “AD-typical” regions. This represents a variant-specific pattern of hypometabolism in addition to a common involvement of temporoparietal cortex across amnestic and non-amnestic variants of AD^20, 41, 42^. This suggests that either the disease epicenter may differ between these AD variants, or that neurodegeneration spreads faster into frontoinsular regions in bvAD compared to tAD, where the frontal regions typically stay spared until more advanced disease stages. As FDG-PET has been suggested to capture the same underlying mechanisms with higher sensitivity than MRI^43^, the overlap of temporoparietal hypometabolic pattern with the atrophy pattern and additional involvement of frontoinsular hypometabolism suggests FDG-PET may capture early spread of neurodegeneration into frontoinsular regions in bvAD. Our findings in a clinically defined group of bvAD patients are in line with a FDG-PET study showing reduced frontal hypometabolism in AD patients with pronounced neuropsychiatric symptoms as indicated by a behavioral questionnaire^44^.

The involvement of the anterior DMN as well as the posterior DMN in bvAD patients provides insights into their clinical presentation, as the anterior DMN is associated with social cognitive functions, such as affective self-referential processing and the inference of other’s mental state^25, 45^, whereas the posterior DMN has been related to several cognitive processes including temporal episodic memory and thinking about the future^25, 46^. bvAD patients resembled tAD patients in the involvement of posterior DMN (reflecting their shared underlying AD pathology or cognitive profile), while bvAD patients showed equivalent involvement of the anterior DMN as bvFTD patients (reflecting their shared behavioral phenotype).

On structural MRI, no subcortical or white matter region was greater affected in bvAD compared to tAD and bvFTD, and the regional WMHV profiles showed more similarities with tAD than with bvFTD. The only deep gray matter structure that showed differences between bvAD and tAD was the amygdala, which showed larger volumes in patients with bvAD than tAD. Several neuropsychiatric disorders have been associated with enlarged amygdalae, such as depression^47^, autism^48^, and carriers of genetic risk variant for panic disorder and anxiety comorbidity in depression^49^. As bvAD patients showed no differences with cognitively normal controls, one speculative explanation might be that patients with bvAD exhibit larger premorbid or baseline amygdalae volumes and, with a similar rate of amygdalar atrophy, progress to the same volumes as cognitively normal individuals. Alternatively, the amygdala may fall further downstream from the pathophysiological epicenter in bvAD and remain more preserved than in tAD. Due to its central role in fear processing and responsivity to emotionally salient stimuli, this structure may be of importance to the clinical phenotype of bvAD.

The ROC analyses showed that functional measures play a pivotal role in the differentiation of bvAD from tAD and bvFTD. These results are in line with previous work investigating the role of FDG-PET in diagnosing bvAD^44^. In addition to neuroimaging markers, there is a need for improving clinical diagnostic tools. As the assessment of presence of behavioral abnormalities currently largely depend on subjective ratings of either clinicians or caregivers, future studies should focus on exploring more objective ways to measure behavioral disturbances, e.g. social cognition test batteries, observations or validated questionnaires in order to improve diagnostic accuracy.

Strengths of the current study include the relatively large sample of clinically defined bvAD patients with multiple neuroimaging markers available. This allowed for a comprehensive examination of neurobiological features and the clinical utility of a broad range of diagnostic tools in this relatively rare variant of AD. The results of the present study also need to be viewed in light of some limitations. First and foremost, data availability across imaging modalities varied. Although we showed that the ROC analyses yielded largely the same results when performed in patients with both MRI and FDG-PET modalities available, this is a major limitation that is due to the retrospective nature of the study as well as the unstandardized data collection. Other limitations include the lack of fMRI data in this group to study functional connectivity.

Overall, somewhat contrary to our hypotheses, bvAD patients showed greater overlap of neuroimaging features with tAD than with bvFTD, further emphasizing their classification as AD patients as opposed to FTD patients with comorbid amyloid pathology^10^. Our results show that differences between bvAD and tAD may lie in functional neuroimaging measures, rather than neuroanatomical measures and may explain, to some extent, the prominent behavioral presentation in bvAD. Indeed, functional measures were strong discriminators in the head-to-head comparison of diagnostic accuracy in the differentiation of bvAD from bvFTD and tAD.

However, future studies should investigate other potential neurobiological factors, such as distribution of tau pathology and involvement of other pathological mechanisms such as decreased Von Economo Neuron^50^ density in the anterior cingulate cortex (associated with social behavior), as well as premorbid personality traits and social cognition in a prospective cohort of bvAD patients, in order to understand the peculiar behavioral presentation in this AD variant that seems to hold relatively little reference to our existing conception of clinico-anatomical relationships.

